# Detection of SARS-CoV-2 in the air from hospitals and closed rooms occupied by COVID-19 patients

**DOI:** 10.1101/2020.12.30.20248890

**Authors:** Shivranjani C Moharir, T. Sharath Chandra, Arushi Goel, Bhuwaneshwar Thakur, Gurpreet Singh Bhalla, Dinesh Kumar, Digvijay Singh Naruka, Ashwani Kumar, Amit Tuli, Swathi Suravaram, Thrilok Chander Bingi, M Srinivas, Rajarao Mesipogu, Krishna Reddy, Sanjeev Khosla, Karthik Bharadwaj Tallapaka, Rakesh K Mishra

## Abstract

To understand air transmission characteristics of SARS-CoV-2 and risks for health care personnel and visitors to hospitals, we analyzed air samples collected from various enclosures in hospitals at Hyderabad and Mohali and performed closed room experiments with COVID-19 positive individuals. We collected 64 air samples from COVID and non-COVID areas of various hospitals and 17 samples from closed rooms occupied by COVID patients. 4 samples from COVID care areas were positive for SARS-CoV-2 with no obvious predilection towards ICU/non-ICU areas in the hospital samples. In the closed room experiments, where one or more COVID-19 patients spent a short duration of time, one sample - collected immediately after the departure of three symptomatic patients from the room - was positive. Our results indicate that the chance of picking up SARS-CoV-2 in the air is directly related to a number of COVID positive cases in the room, their symptomatic status, and the duration of exposure and that the demarcation of hospital areas into COVID and non-COVID areas is a successful strategy to prevent cross infections. In neutral environmental conditions, the virus does not seem to spread farther away from the patients, especially if they are asymptomatic, giving an objective evidence for the effectiveness of physical distancing in curbing the spread of the epidemic.

## Introduction

It is astonishing how a ∼100nm viral particle, SARS-CoV-2 (severe acute respiratory syndrome coronavirus-2) has affected the human life in the most unexpected ways in just a few months [1]. The effects on the patient are seen to range from being totally asymptomatic to respiratory or multiorgan failure and the complications are also not just restricted to respiratory system [2]. In spite of the multiple measures taken by countries globally, containing the virus has been challenging. Initially, contact and droplet-mediated transmission were considered as major modes of transmission for the SARS-CoV-2. Accordingly, hand washing and social distancing were the main measures suggested along with wearing masks, to avoid contracting the disease. The alarmingly increasing number of cases of COVID-19 globally raised the possibility of airborne transmission of SARS-CoV-2 [3]. The antecedent of SARS-CoV-2, SARS-CoV-1, which had an outbreak in 2002-2004, was reported to spread through air [4, 5] and through viral aerosols generated by patients [6]. Apart from SARS-CoV-1, other viruses - Norwalk like virus [7], respiratory syncytial virus [8], MERS (Middle East Respiratory Syndrome) coronavirus [9] and Influenza A/H591 virus between ferrets [10] were also reported to have the possibility of airborne transmission. Considering that these viruses, especially SARS-CoV-1 and MERS virus, which are closely related to SARS-CoV-2, were capable of getting transmitted through air, the possibility of airborne transmission of SARS-CoV-2 cannot be ruled out [3, 11]. More so, when it has been observed that SARS-CoV-2 is quite stable in aerosols [12] and that it is more stable in aerosols when compared to SARS-CoV-1 and MERS [13].

There are existing reports of surface and environment contamination with SARS-CoV-2 in hospital rooms of COVID-19 positive patients [14-16]. A study by Lednicky et al. (2020) has provided evidence for the presence of viable SARS-CoV-2 in the air samples collected from hospital room with COVID-19 patient even in the absence of any aerosol generating procedure [17]. Rami et al. (2020) also suggested that, in hospitals, droplets containing SARS-CoV-2, with strong directional airflow, can spread the virus farther than 2 meters [18]. To get further insights on transmission characteristics of SARS-CoV-2 in air and to assess the risk for healthcare workers, we collected air samples from different locations of hospitals and from closed rooms in which either one or more COVID positive individuals were present during sampling.

## Materials and methods

### Hospital settings

Air samples were collected from three hospitals from Hyderabad, India - Hospital 1, Hospital 2 and Hospital 3; and three other hospitals from Mohali, India –Hospital 4, Hospital 5 and Hospital 6. Air samples were collected from COVID ICUs, nurse stations, COVID wards, corridors, non-COVID wards, PPE doffing areas, COVID rooms, OP corridors, mortuary, COVID casualty areas, non-COVID ICUs and doctors’ rooms. The details of air sampling conditions are mentioned in Tables 2 & 3 and Supplementary Tables 1 & 2.

**Table 1:** Air samples collected from hospitals in two cities in India-Hyderabad and Mohali showing the presence of SARS-CoV-2 in air: 64 air samples from different locations of 3 hospitals each from two cities in India-Hyderabad and Mohali, were analyzed for the presence of SARS-CoV-2. 4 out of 64 samples were positive for SARS-CoV-2 and 16 others showed traces of the virus. The air samples were considered ‘positive’ when they satisfied positivity criteria as per the RT-PCR kit’s specifications for clinical nasopharyngeal swab samples. Air samples were denoted as ‘showing traces of SARS-CoV-2 RNA’ when they did not fit the positivity criteria, but showed the following trends: either amplification of one of the SARS-CoV-2 viral genes in at least one of the triplicates or Ct values higher than the prescribed cut-offs (Ct> 36).

| City | Total no. of samples | No. of positive samples | No. of samples showing traces of SARS-CoV-2 RNA |
| --- | --- | --- | --- |
| Hyderabad | 41 | 3 | 7 |
| Mohali | 23 | 1 | 9 |
| **Total** | **64** | **4** | **16** |

**Table 2:** Analysis for SARS-CoV-2 presence at various locations of the 3 hospitals in Hyderabad. A total of 41 air samples were collected from indicated locations of 3 hospitals. The air samples were considered ‘positive’ when they satisfied positivity criteria as per the RT-PCR kit’s specifications for clinical nasopharyngeal swab samples. Air samples were denoted as ‘showing traces of SARS-CoV-2 RNA’ when they did not fit the positivity criteria, but showed the following trends: either amplification of one of the SARS-CoV-2 viral genes in at least one of the triplicates or Ct values higher than the prescribed cut-offs (Ct> 36). UD: Undetectable

| Hospital | Locations of sample collection | Air Sample Ct Value |  |  | Interpretation |
| --- | --- | --- | --- | --- | --- |
|  |  | E Gene | N Gene | ORF1ab |  |
| Hospital 1 | COVID ICU 1 | 32.78 | 30.54 | 30.51 | Positive |
|  | COVID ICU 2 | UD | UD | UD |  |
|  | Nurse Station | UD | UD | UD |  |
|  | COVID Ward | 30.47 | 28.51 | 29.03 | Positive |
|  | Corridor | 34.10 | UD | 31.75 | Showing traces |
| Hospital 2 | Non-COVID Ward 1 | UD | UD | UD |  |
|  | Non-COVID Ward 2 | UD | UD | UD |  |
|  | COVID Nurse Station | UD | UD | UD |  |
|  | COVID ICU 1 | UD | UD | UD |  |
|  | COVID ICU 2 | UD | UD | UD |  |
|  | PPE Doffing Area | UD | UD | UD |  |
|  | COVID Room | UD | UD | UD |  |
|  | OP Corridor | UD | UD | UD |  |
| Hospital 3: Sampling 1 | Non-COVID Ward | UD | UD | UD |  |
|  | COVID Nurse Station | 33.20 | 29.82 | 31.96 | Positive |
|  | Mortuary | UD | 33.84 | UD | Showing traces |
|  | OP Corridor | UD | UD | UD |  |
|  | COVID Casualty | UD | UD | UD |  |
|  | PPE Doffing Area | UD | UD | UD |  |
|  | Non-COVID ICU | UD | UD | UD |  |
|  | COVID Room | UD | UD | UD |  |
|  | COVID ICU | UD | UD | UD |  |
| Hospital 3: Sampling 2 | OP Corridor | UD | 28.73 | UD | Showing traces |
|  | Mortury | UD | UD | UD |  |
|  | COVID Casualty | UD | UD | UD |  |
|  | COVID ICU | UD | 30.68 | 37.10 | Showing traces |
|  | COVID Nurse Station | UD | UD | UD |  |
|  | PPE Doffing Area | UD | UD | UD |  |
|  | Non-COVID ICU | UD | UD | UD |  |
| Hospital 3: Sampling 3 | OP Corridor | UD | UD | UD |  |
|  | Mortuary | UD | UD | UD |  |
|  | Non-COVID ICU | UD | UD | UD |  |
|  | COVID ICU | UD | 31.57 | UD | Showing traces |
|  | COVID Nurse Station | UD | UD | UD |  |
|  | PPE Doffing Area | UD | UD | UD |  |
| Hospital 3: Sampling 4 | COVID Room 1 Sample 1 | UD | UD | UD |  |
|  | COVID Room 1 Sample 2 | UD | UD | UD |  |
|  | COVID Room 1 Sample 3 | UD | 32.00 | UD | Showing traces |
|  | COVID Room 2 Sample 1 | UD | UD | UD |  |
|  | COVID Room 2 Sample 2 | UD | 32.08 | UD | Showing traces |
|  | COVID Room 2 Sample 3 | UD | UD | UD |  |

### Closed-room controlled experiments

Asymptomatic or mildly symptomatic COVID-19 positive individuals were asked to sit in a closed room during air sample collection procedure without masks and were allowed to talk over phone or interact with each other. Immediately after sample collection procedure was over, they were allowed to leave the room. Air samples from the closed room were collected prior to arrival of the COVID-19 positive individuals, from distances between 4-12 feet from the individual, immediately after their departure and 2-6 hours post their departure. The pre-arrival and post departure samples were collected from the place where the participants sat in the room. The details of air sampling conditions are mentioned in Table 4.

### Swab sample collection

For closed-room experiments, nasopharyngeal swabs were collected in viral transfer medium (VTM) from the participants before air sampling (could not be done for one individual). The swabs were processed according to standard procedures and SARS-CoV-2 viral RNA Ct values were determined.

### Air sample collection

Air samples were collected on disposable gelatin filters (Sartorius, Cat. No. 17528-80-ACD) using AirPort MD8 air sampler (Sartorius, Cat. No. 16757). 1000 liters of air was collected at a flow rate of 50 liters per minute and a sampling time of 20 minutes. After sample collection, the gelatin membrane was aseptically removed, dissolved in 4 ml of Tris-EDTA buffer (10mM Tris and 0.1mM EDTA; pH 7.4) or VTM at 37°C in a water bath for 10 minutes or till it completely dissolved and then used for RNA extraction. (exception - samples collected during sampling 1 at Hospital 4 and 5 in Mohali, were collected using Whatman filter paper placed on 90 mm petri dish using Merck’s MAS-100® series air sampler, 1000 L at a rate of 100 liters per minute).

### RNA extraction and Real Time-RT-PCR

1ml sample containing dissolved gelatin membrane was used for RNA extraction by guanidinium thiocyanate-phenol-chloroform extraction-column method. The sample was lysed using QIAzol Lysis Reagent (Qiagen, Cat. No. 79306) and incubated at room temperature for 10 minutes. After adding chloroform, mixing and centrifuging at 12,000 X g for 10 minutes at 4°C, the RNA containing aqueous layer was used for RNA isolation using QIAamp Viral RNA Mini kit (Qiagen; Cat No. ID: 52906) according to manufacturer’s protocol. The extracted RNA was used for SARS-CoV-2 E gene, N gene and ORF1ab gene detection using Fosun COVID-19 RT-PCR Detection Kit (Shanghai Fosun Long March Medical Science Co., Ltd; Cat. No. PCSYHF03-a). RT-PCR was run on QuantStudio™ 5 Real Time PCR system. RT-PCR was done in triplicates for each sample and data was analysed using Design and Analysis Software v1.5.1 QuantStudio™ 5. For sampling 1 of Hospital 4 and 5 at Mohali, RNA was isolated using AURA PURE Viral RNA Isolation Kit (Aura Biotechnologies Private Ltd., Cat. No. MNP-R004-100) and RT-PCR was performed using TRUPCR^®^ COVID-19 Real-Time RT-PCR Kit (3B Blackbio Biotech India Ltd; Cat No. 3B304).

The air samples were considered ‘positive’ when they satisfied positivity criteria as per the RT-PCR kit’s specifications for clinical nasopharyngeal swab samples. Air samples were denoted as ‘showing traces of SARS-CoV-2 RNA’ when they did not fit the positivity criteria, but showed the following trends: either amplification of one of the SARS-CoV-2 viral genes in at least one of the triplicates or Ct values higher than the prescribed cut-offs (Ct> 36).

## Results

### SARS-CoV-2 is detected in a few hospital air samples

We analyzed a total of 64 air samples from various locations at hospitals from two cities in India-Hyderabad and Mohali. Out of the 64 samples collected, 4 samples collected from COVID care areas were positive for SARS-CoV-2 (Table 1).

From Hyderabad, 41 air samples were collected from 3 hospitals between September 2020 and November 2020. Hospital 1 is a large tertiary care government hospital, being exclusively used for COVID care at the time of sample collection. Hospital 2 is a private trust hospital catering to very limited number of COVID patients while Hospital 3 is again a tertiary care government set-up catering to a moderate number of COVID patients along with other medical services. Of all the places tested, samples collected from an ICU and a COVID general ward in Hospital 1 and a nursing station in Hospital 3 were positive (Table 2). From Mohali, 23 air samples were collected from 3 hospitals (all tertiary care hospitals catering to moderate number of COVID and non COVID patients) between July and December. Among these, only one ICU sample in Hospital 5 was positive (Table 3).

**Table 3:**
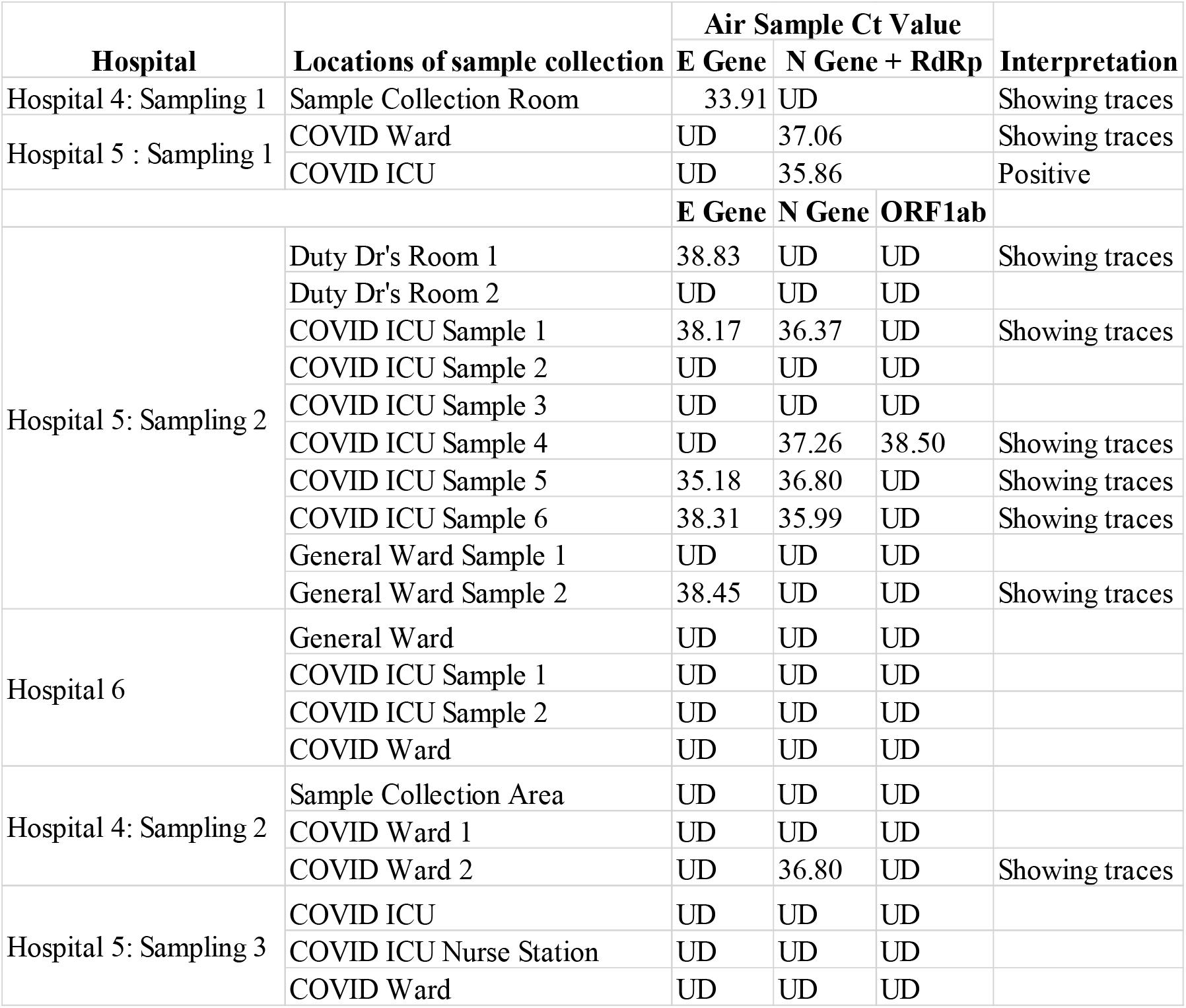
Analysis of SARS-CoV-2 at various locations of 3 hospitals in Mohali. A total of 23 air samples were collected from indicated locations of 3 hospitals. The air samples were considered ‘positive’ when they satisfied positivity criteria as per the RT-PCR kit’s specifications for clinical nasopharyngeal swab samples. Air samples were denoted as ‘showing traces of SARS-CoV-2 RNA’ when they did not fit the positivity criteria, but showed the following trends: either amplification of one of the SARS-CoV-2 viral genes in at least one of the triplicates or Ct values higher than the prescribed cut-offs (Ct> 36). For samples collected during sampling 1 of Hospital 4 and 5, True PCR kit which can detect SARS-CoV-2 E gene and N gene/ RdRp was used for RTPCR. For other samples, Fosun COVID-19 RT-PCR Detection Kit which detects SARS-CoV-2 E gene, N gene and ORF1ab gene was used. UD: Undetectable

**Table 4:** Analysis of air samples from closed room occupied by COVID-19 positive individuals. 7 COVID-19 positive individuals were asked to spend indicated time in a closed room during air sample collection. Each sampling was performed for 20 minutes. Air samples were collected at the indicated distances from the individuals and analyzed for the presence of SARS-CoV-2. The air samples were considered ‘positive’ when they satisfied positivity criteria as per the RT-PCR kit’s specifications for clinical nasopharyngeal swab samples. Air samples were denoted as ‘showing traces of SARS-CoV-2 RNA’ when they did not fit the positivity criteria, but showed the following trends: either amplification of one of the SARS-CoV-2 viral genes in at least one of the triplicates or Ct values higher than the prescribed cut-offs (Ct> 36). UD: Undetectable

| Expt No. | No. of Individuals in room | Symptoms | Swab Ct value | Distance from individual | Air sample Ct value |  |  | Interpretation | Time spent |
| --- | --- | --- | --- | --- | --- | --- | --- | --- | --- |
|  |  |  |  |  | E Gene | N Gene | ORF1ab |  |  |
| 1 | 1 | Asymptomatic | 28.87 | 4 feet | UD | UD | UD |  | Sampling time |
|  |  |  |  | 12 feet | UD | UD | UD |  | 20 minutes + Sampling time |
| 2 | 3 | Asymptomatic | 28.87, 31.76, 32.47 | 4 feet | UD | UD | 32.12 | Showing traces | Sampling time |
|  |  |  |  | 12 feet | UD | UD | UD |  | 20 minutes + Sampling time |
|  |  |  |  | Pre-Arrival | UD | UD | UD |  | 0 minutes |
|  |  |  |  | Departure | UD | 32.4 | UD | Showing traces | 40 minutes |
|  |  |  |  | Post-departure 2-6 hrs | UD | UD | 32.26 | Showing traces | 40 minutes |
| 3 | 1 | Asymptomatic | 27.57 | 4 feet | UD | UD | UD |  | Sampling time |
|  |  |  |  | 8 feet | UD | UD | UD |  | 20 minutes + Sampling time |
|  |  |  |  | 12 feet | UD | UD | UD |  | 40 minutes + Sampling time |
|  |  |  |  | Departure | UD | UD | UD |  | 60 minutes |
| 4 | 3 | Mild symptoms | 29.1, 19.85, N* | 4 feet | UD | UD | UD |  | Sampling time |
|  |  |  |  | 8 feet | UD | UD | UD |  | 20 minutes + Sampling time |
|  |  |  |  | 12 feet | UD | UD | UD |  | 40 minutes + Sampling time |
|  |  |  |  | Pre-Arrival | UD | UD | UD |  | 0 minutes |
|  |  |  |  | Departure | 34.468 | 31.017 | UD | Positive | 60 minutes |
|  |  |  |  | Post-departure 2-6 hrs | UD | 33.349 | UD | Showing traces | 60 minutes |

### SARS-CoV-2 detection in closed room experiments

To understand how far and for how long SARS-CoV-2 can be detected in the air, when COVID-19 positive individuals spend time in a closed room, we analyzed air samples at different distances from COVID-19 positive individuals and at different time points. The participants were made to sit in one corner of a room with no perceived air flow for short span of time and air samples were collected at varying distances from them. A positive result was obtained only in the immediate post departure sample from the place they were sitting in experiment 4 where 3 mildly affected individuals had left the room after staying for about an hour (Table 4).

## Discussion

In the initial phases of the pandemic, the transmission of SARS-CoV-2 was largely thought to be through contact and droplet spread. However, with studies reporting transmission among physically distanced individuals in closed spaces with air conditioning [18, 19] and the fact that, the viral spread could not be effectively curbed in spite of strict lockdowns in various countries of the world, raised the possibility of its airborne transmission. CDC also released a statement acknowledging the possibility of air borne transmission in certain scenarios (https://www.cdc.gov/coronavirus/2019-ncov/more/scientific-brief-sars-cov-2.html).

In our study, from the air samples collected in hospitals, virus could be detected from various COVID care areas with no specific predilection towards ICU/ non-ICU areas. The virus could not be detected in any of the non-COVID areas, providing objective evidence that the strategy of separating hospital premises into COVID and non-COVID care areas is effective. The positivity rate was found to be higher when the number of COVID patients were higher in the room, a finding concordant with results of closed room experiments as well. A point to be highlighted from the hospital experiments was that in 3/4 samples which were positive, the sampler was at least 10 feet away from the nearest patient. As there is no record on events that occurred in the sampling area before the sample collection began, we are unable to infer this finding. But this may be an indicator that long term presence of COVID positive patients in an enclosed space may contribute to a significant increase in aerosol burden in the air.

In our study, we found that the virus does not travel much in the air in neutral environmental conditions (ambient temperature and humidity with no perceived air flow), especially if duration of exposure is short. Virus could not be picked up at a distance of even 4 feet when COVID positive individuals spent a short time (20 minutes) in the room. This indicates that short duration of exposure to a COVID positive individual may not put one at a significantly increased risk. The samples collected at 8 feet and 12 feet subsequently were also negative. In experiment 4, where three people with mild symptoms, were in a room for at least an hour, virus could be detected at the same place immediately after their departure. A finding strengthening the logic that chances of finding the virus in air are more when more number of symptomatic people stay in a closed place for longer periods.

These findings have significant implications in the current situation, when many countries have relaxed the restrictions on public mobility and interactions, even when the number of COVID-19 cases are increasing steadily. In many densely populated nations where the recommended physical distancing norms may be difficult to implement in public/ office spaces, distancing as much as possible with usage of masks should be actively promoted. Whilst, any form of verbal communication from closed quarters without wearing masks should be prohibited.

The strict lockdowns implemented in countries like India have brought the economy to a standstill. We are in a phase of recovery and possibly cannot afford any other such economic disruptions. On one hand, a sense of disbelief/ disregard for the advocated preventive measures has actually set in the people of our country, as the cases continued to spike in spite of such strict lockdown. On the other hand, there is a large section of the population who have not returned back to their jobs due to the fear of contracting the disease. It is evident that the spread of the pandemic may be largely attributed to noncompliance of COVID-19 safety guidelines on the part of the people. A right amount of caution with courage backed up by sound scientific principles is the need of the hour. The findings of our study objectively reassure people that advocated preventive measures would largely be successful in preventing the infection and urge the governments to continue promoting the same. Short-term travel and resumption of economic activity can be safe if adequate measures are taken.

However, it is to be noted that many of the air samples from hospitals and closed room experiments showed PCR signal for one of the SARS-CoV-2 genes or had Ct values above the prescribed cut-off. The possibility that SARS-CoV-2 is actually present in these air samples cannot be ruled out completely considering the diluting effect due to diffusion of the virus in air and the fact only 1000 liters (1 cubic meter) of air was sampled each time. If we consider these results as an evidence for presences of traces of viral RNA but not as negatives or false positives, all the propositions made in the paper so far still hold true except that the virus could be detected in the air long after the patients had left the room. This interpretation along with the results suggesting the presence of virus at greater than 10 feet from the patients in the hospital samples, in fact are supportive of air borne transmission. Further experiments are required not only to affirm/ negate this line of thought but also to establish the infectivity of such positive air samples.

## Data Availability

all data is available

## Acknowledgements

The authors thank the COVID-19 positive individuals who allowed us to take air samples from their vicinity. The authors acknowledge the help of Amareshwar Vodapalli and S Reddy Mahesh in conducting the experiments.

## Ethical Approval

The study was approved by the Institutional Ethics Committee of CSIR-Centre for Cellular and Molecular Biology (IEC-83/2020).

---

**Supplementary table 1:**
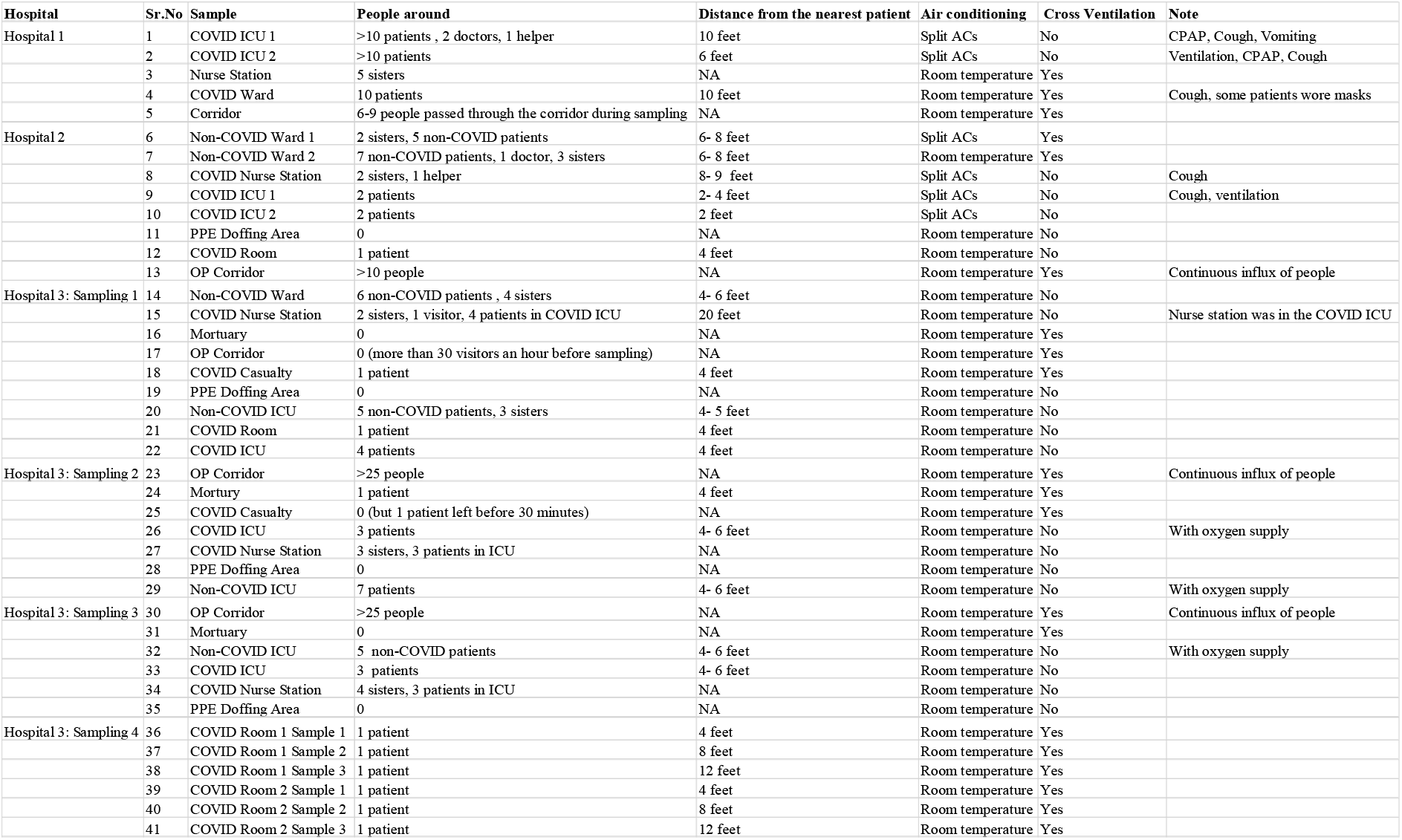
Details of air samples collected from hospitals in Hyderabad.

**Supplementary table 2:**
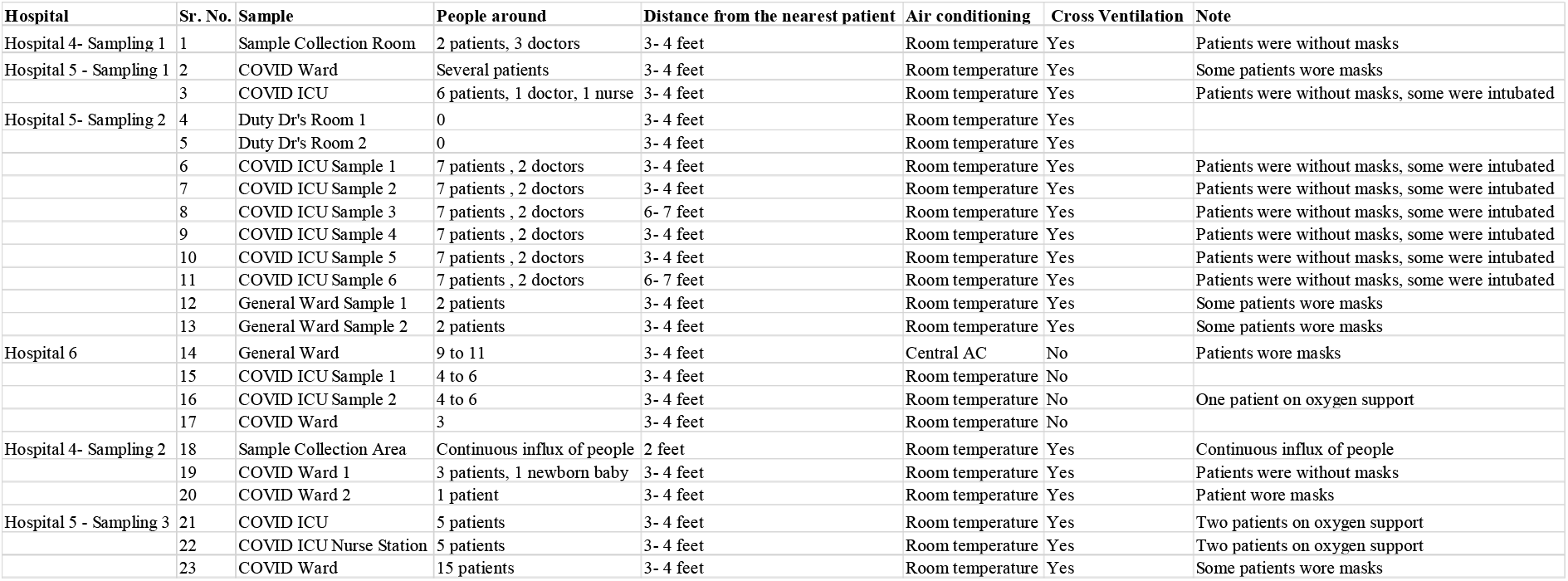
Details of air samples collected from hospitals in Mohali.

